# A novel disease mechanism leading to the expression of a disallowed gene in the pancreatic beta-cell identified by non-coding, regulatory mutations controlling *HK1*

**DOI:** 10.1101/2021.12.03.21267240

**Authors:** Matthew N. Wakeling, Nick D. L. Owens, Jessica R. Hopkinson, Matthew B. Johnson, Jayne A.L. Houghton, Antonia Dastamani, Christine S. Flaxman, Rebecca C. Wyatt, Thomas I. Hewat, Jasmin J. Hopkins, Thomas W. Laver, Rachel Van Heugten, Michael N. Weedon, Elisa De Franco, Kashyap A. Patel, Sian Ellard, Noel G. Morgan, Edmund Cheesman, Indraneel Banerjee, Andrew T. Hattersley, Mark J. Dunne, International Congenital Hyperinsulinism Consortium, Sarah J. Richardson, Sarah E. Flanagan

## Abstract

Gene expression is tightly regulated with many genes exhibiting cell-specific silencing when their protein product would disrupt normal cellular function. This silencing is largely controlled by non-coding elements and their disruption might cause human disease. We performed gene-agnostic screening of the non-coding regions to discover new molecular causes of congenital hyperinsulinism. This identified 14 non-coding *de novo* mutations affecting a 42bp conserved region encompassed by a regulatory element in intron 2 of *Hexokinase 1* (*HK1*), a pancreatic beta-cell disallowed gene. We demonstrated that these mutations resulted in expression of *HK1* in the pancreatic beta-cells causing inappropriate insulin secretion and congenital hyperinsulinism. These mutations identify a regulatory region critical for cell-specific silencing. Importantly, this has revealed a new disease mechanism for non-coding mutations that cause inappropriate expression of a disallowed gene.

---

Genetic discovery in Mendelian disease has focused on identifying highly penetrant mutations affecting the function of genes expressed in clinically affected tissue(s). Whilst this approach has proven successful, the underlying aetiology of over 3000 presumed monogenic diseases remains undefined^1-4^, of which many have significant clinical and genetic heterogeneity^5,6^. Congenital Hyperinsulinism (CHI), is characterised by inappropriate insulin secretion during hypoglycaemia. It is a clinically and genetically heterogeneous disease where, despite extensive sequencing efforts, the underlying aetiology is not known in up to 50% of individuals^7,8^.

To identify new aetiologies, we performed whole-genome sequencing (WGS) on 135 individuals with biochemically confirmed, persistent CHI without an identified mutation in a known disease causing gene (**Supplemental Table 1**). We initially searched for genes containing novel coding *de novo* or biallelic variants in two or more individuals. When no such genes were found, we turned to the non-coding genome and searched for *de novo* copy number variants. This identified a single locus within intron 2 of Hexokinase 1 (*HK1*) containing two heterozygous ∼4.5Kb deletions with a 2,787bp overlap in two unrelated individuals (**Supplemental Figure 1**). Digital droplet PCR confirmed the deletions in both probands and an affected twin, in keeping with a germline mosaic mutation.

We next searched for *de novo* single nucleotide variants and indels within the minimal deleted region within our WGS discovery cohort and in a replication cohort of 27 individuals with CHI who had undergone pancreatectomy as part of routine clinical care (**Supplemental Table 1**). This identified 7 different *de novo* mutations in 12 probands. In two cases the mutation had been inherited by similarly affected offspring (**Figure 1**). All mutations were novel^9,10^ and within a 42bp conserved region constrained against variation in gnomAD.^11^

**Figure 1.**
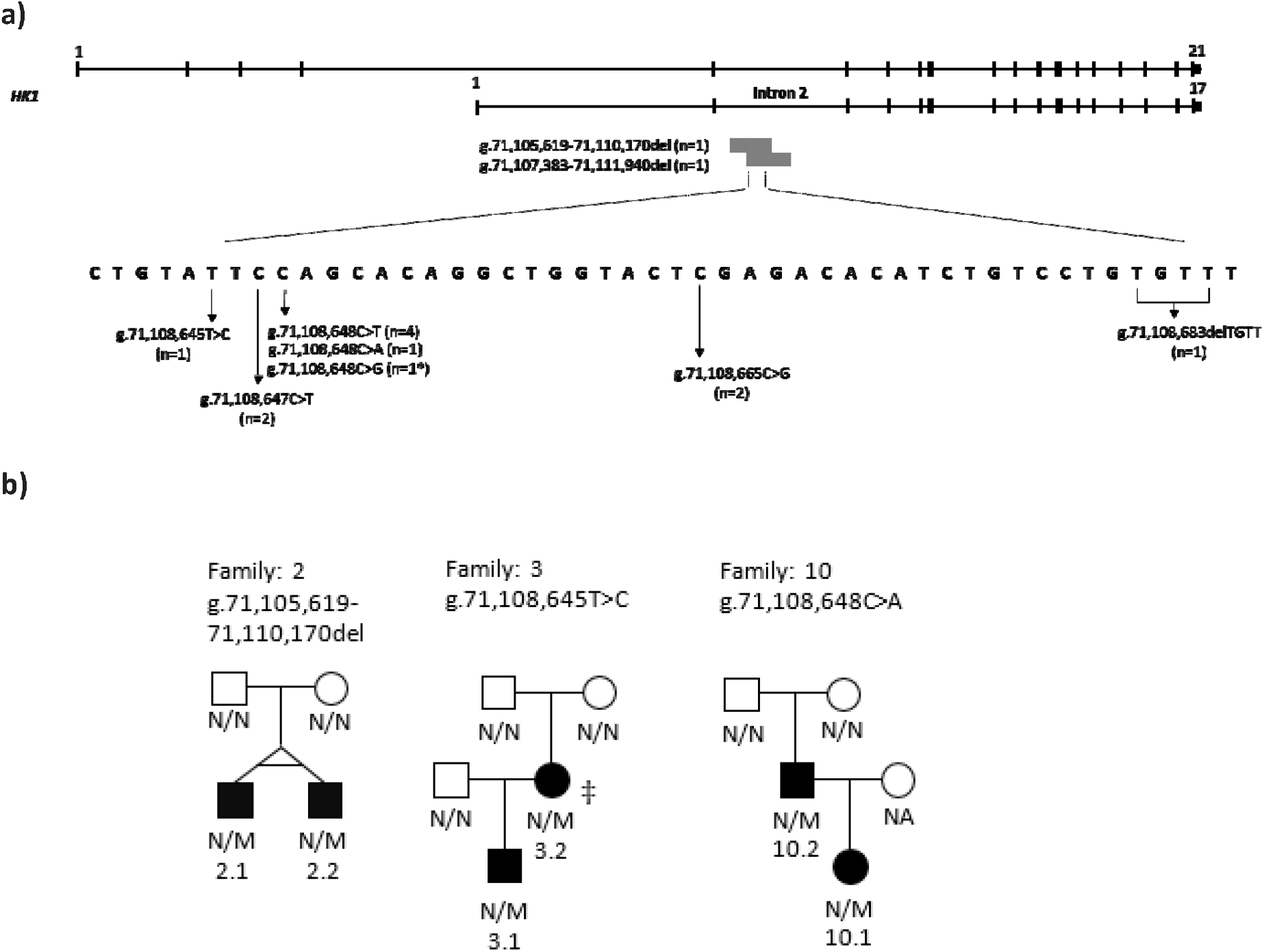
**a**) Schematic representation of the *HK1* gene (GRCh37/hg19 chr10:71,029,756-71,161,637). The full-length testis-specific isoform (ENST00000448642) and the shorter ubiquitously-expressed isoform (ENST00000359426) are depicted. The positions of the two ∼4.5Kb deletions within intron 2 of the shorter isoform and the 7 different heterozygous point mutations/indels within the minimal deleted region identified in 14 probands are shown. The number of probands with each mutation is provided. An asterisk (*) denotes a single proband with two *de novo* mutations (see **Supplementary Figure 7**). The mutation positions are given according to the GRCh37/hg19 genomic coordinates. b) Partial pedigrees showing inherited *HK1* mutations in 3 families. Filled symbols represent individuals with congenital hyperinsulinism. (‡) Haplotype analysis confirmed that the *HK1* mutation had arisen *de novo* in patient 3.2. M, *HK1* mutation; N, no mutation; NA, DNA not available. Pedigrees for the 11 probands with *de novo HK1* mutations are not shown.

Overall, the finding of 9 different *de novo* mutations in 14 probands, co-segregating with disease in three additional family members provides overwhelming evidence of disease-causality.

All 17 individuals with a *HK1* non-coding mutation had severe early-onset CHI (median age at diagnosis: Birth [IQR: 0-14 days]). These mutations were not associated with macrosomia at birth (median birthweight z score: 0.61 [IQR: 0.10-1.84]) suggesting that insulin secretion was not markedly increased *in utero*. In all cases the hyperinsulinism persisted, with the eldest individual still known to be affected being 18 years of age. These findings suggest that the mutations act to disrupt glucose-induced insulin secretion throughout post-natal life and into adulthood but have less impact during fetal development.

The *HK1* mutations caused a beta-cell specific defect with no common additional features between patients. In five individuals, medical management was ineffective, leading to pancreatic resection. Of these, the only case that had resolution of the CHI developed insulin-dependent diabetes following near-total pancreatectomy. Histopathological analysis of resected pancreatic tissue (n=2) demonstrated a discrete pathology, distinguishable from diffuse disease resulting from *ABCC8* mutations, the commonest cause of CHI (**Supplemental Figure 2**). An overview of the clinical features is provided in **Supplemental Table 2**.

*HK1* encodes a glycolytic enzyme that is silenced in the pancreas and liver but expressed in all other mature tissues (**Supplemental Figure 3a-c**) where it supports glucose metabolism, ensuring cell survival^12^. Within the pancreatic beta-cell, glucose is phosphorylated to glucose-6-phosphate by Glucokinase (GCK; Hexokinase 4) which, due to a low-binding affinity (Km ∼8mM) acts as the pancreatic glucose-sensor coupling insulin release to the prevailing glucose concentration^13^. Hexokinase 1 has a markedly higher affinity for glucose (Km <50μM)^13^. Silencing of *HK1* in favour of *GCK* in the beta-cell therefore ensures appropriate glucose-sensing, minimising insulin release at low glucose levels. Whilst bi-allelic loss-of-function *HK1* mutations have been reported to cause non-spherocytic haemolytic anaemia^14^, dominant or recessive coding mutations have not been described in individuals with defects in glucose homeostasis; in keeping with the absence of *HK1* protein in beta-cells.

In resected pancreatic tissue of individuals with non-coding mutations, we found that *HK1* was expressed and co-localised with insulin in the islets of affected tissue but not in unaffected controls (**Figure 2**). *HK1* did not co-localise with glucagon (secreted by pancreatic alpha-cells) suggesting that the impact of the mutations was beta-cell specific (**Supplemental Figure 4**). These results confirmed that the mutations cause *HK1* to be inappropriately expressed in the pancreatic beta-cell and explain why there is increased insulin secretion in these individuals during hypoglycaemia.

**Figure 2:**
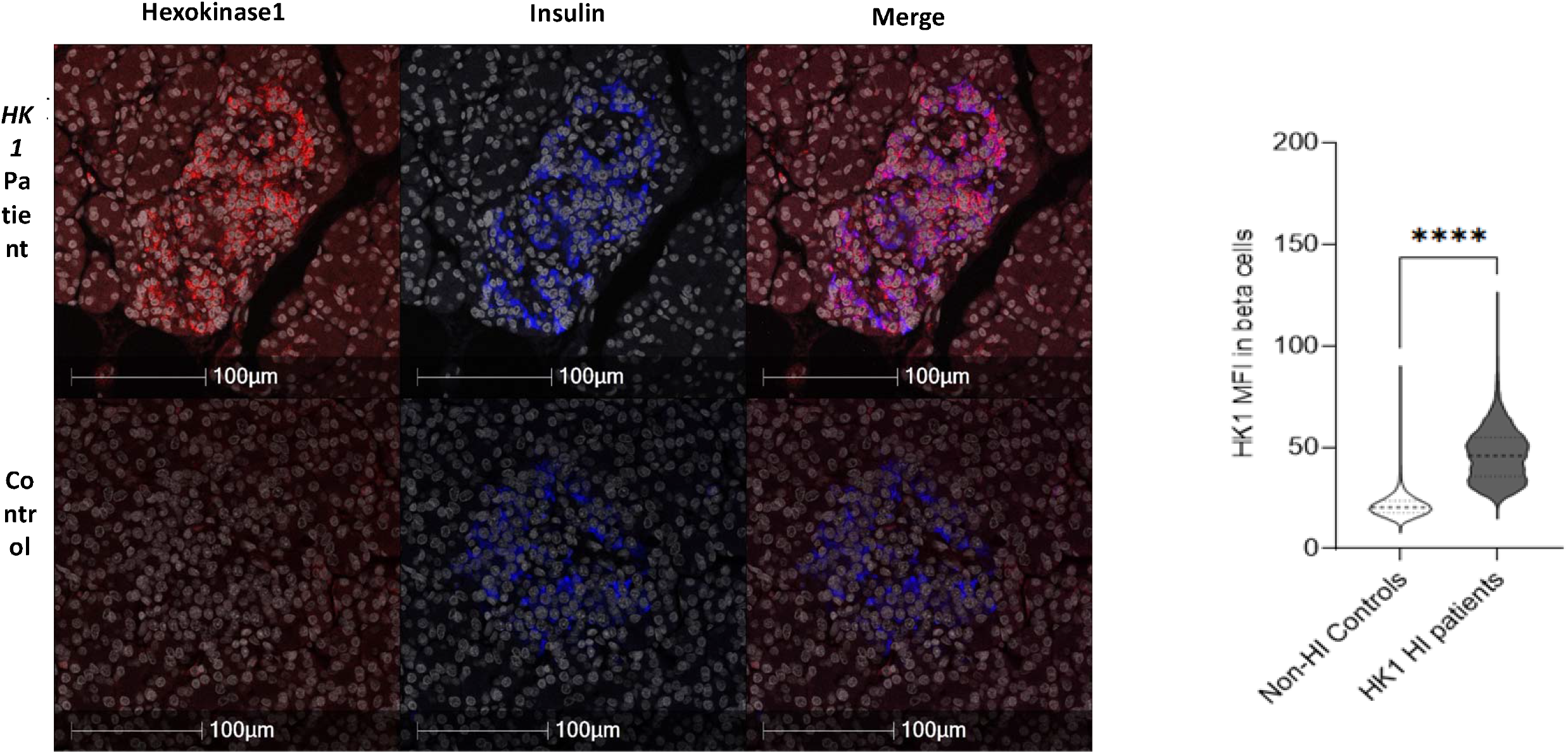
*HK1* expression is present in the beta-cells of donors with *HK1*-CHI, but not controls. **a**) Staining of HK1 (red); insulin (blue) and DAPI (white) in pancreatic tissue resected from one patient with a *HK1* mutation. Within this section HK1co-localises with insulin (purple staining) whereas, no *HK1* expression is observed in the non-hyperinsulinism (Non-HI) control donor. Scale bar – 100μm. **b**) HK1 expression (Median fluorescence intensity (MFI)) is significantly increased in the *HK1* HI donors (n=2 donors; 17015 beta-cells), when compared to Non-HI control donors (n=2 donors; 21408 beta-cells). Two-tailed Mann-Whitney (U=590830; ****p<0.0001).

We identified an islet enhancer encompassing our critical region^15^ bound by a broad set of islet transcription factors, with NKX2-2 and FOXA2 most prominent (**Figure 3**). Single-nuclei ATAC-seq (snATAC-seq) data in human islets^16^ revealed a peak of open chromatin in beta-cells and showed that the relevant *HK1* promoter remains accessible (**Figure 3b**). Human islet Hi-C data^17^ confirmed that both our region and the relevant *HK1* promoter are contained within a well-insulated domain, without evidence of contact to distal loci (**Figure 3d**) further supporting direct regulation by the critical region on the promoter.

**Figure 3:**
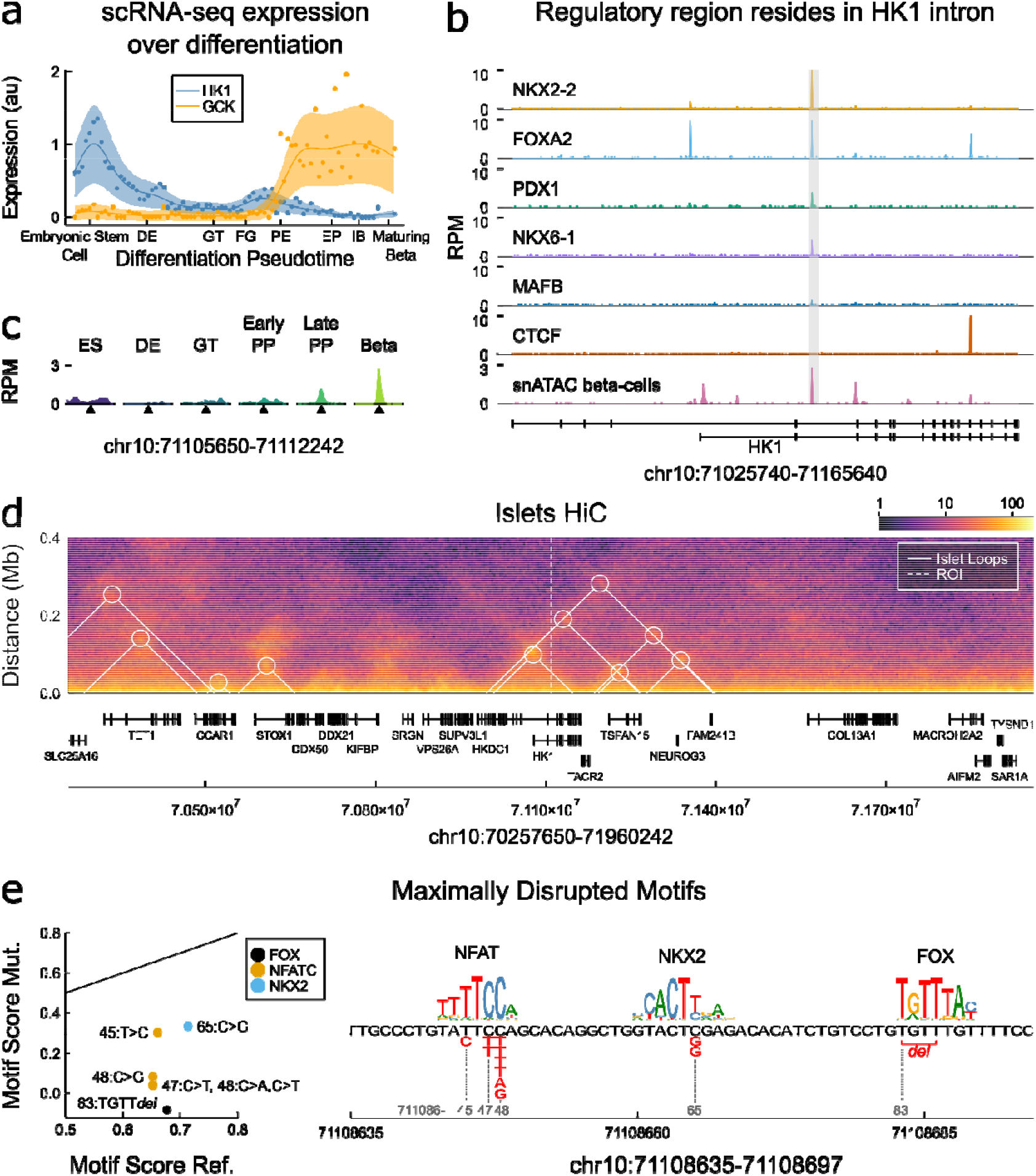
**a**) Expression of *HK1* and GCK for comparison over the course of pancreatic differentiation from embryonic stem cells to maturing beta-cells, expression data over a beta-cell differentiation pseudotime^18^, Gaussian process regression median and 95% CI shown. DE – definitive endoderm, GT – gut tube, FG – foregut, PE – pancreatic endoderm, EP – endocrine precursors, IB – immature beta-cells. See **Supplemental Figure 3** for further *HK1* expression. **b**) Transcription factor binding in human islets^15^ and chromatin accessibility from snATAC-seq in the cluster of cells assigned to beta-cells^16^ over *HK1* locus, the region containing the mutations is marked as grey box. Chromatin accessibility in other endocrine cell types is provided in **Supplemental Figure 5. c**) Chromatin accessibility during pancreatic differentiation over critical region, stages: ES – embryonic stem cell, DE – definitive endoderm, GT – gut tube, Early/Late PP – pancreatic progenitor, describe ATAC-seq from in vitro differentiation. Beta – gives accessibility in snATAC-seq^16^ (as shown in B). **d)** HiC data in human islets^17^ reveals *HK1* is contained within a well-insulated domain, heatmap gives log scale contact frequencies, white triangle and circles mark chromatin loops called in same study. **e)** Transcription factor motif families disrupted by the mutations, shown are those motif families maximally disrupted by each mutation (red font). Notable secondary motif families given in **Supplemental Figure 7 and Supplemental Table 4**. Left shows scatter of normalised motif scores for the reference versus mutated score (motif score normalised by maximal motif score), black line gives 1-1 line of equal motif. Number gives final two digits of chromatin position, e.g. 45 – 71108645. Right gives sequence logo for the three maximally disrupted families. Members of each family are expressed in beta-cells **Supplemental Figure 6**.

During development, *HK1* is highly expressed in embryonic stem cells but is progressively downregulated during pancreatic cell-type differentiation becoming extinguished during beta-cell maturation^18,19^ (**Figure 3a, Supplemental Figure 3d**). GCK shows increased expression during differentiation as *HK1* is downregulated (**Figure 3a**). Despite being absent from beta-cells *HK1* is expressed in mature stellate- and duct-cells suggesting that it becomes de-repressed in other pancreatic cell-types (**Supplemental Figure 3e**).

Chromatin accessibility data shows our critical region remains closed over pancreatic-cell differentiation until pancreatic progenitor stages, when it becomes accessible and bound by a complement of factors in the pancreatic progenitor regulatory network^20,21^ (**Figure 3c, Supplemental Figure 5b-c**). This suggests the region is necessary for HK1 repression in late pancreatic development and mediating beta-cell specific control in adult cells but that it is not required for HK1 repression during early differentiation.

Analysis of histone modifications during pancreatic differentiation, in human islets^20^ and the EndoC-βH1 beta-cell line^22^, revealed that repression is actively maintained. The critical region is marked by a bivalent state encompassed by focal peaks of active enhancer marks H3K4me1 and H3K27ac and a broader polycomb H3K27me3 domain (**Supplemental Figure 5d**).

Analysis of transcription factor motif families revealed that mutations disrupt transcription factor binding sites for FOX, NKX2 and NFAT families (**Supplemental Table 4, Figure 3c**), which each have family members expressed in beta-cells (**Supplemental Figure 6**). This is supported by ChIP-seq data revealing that NKX2-2 and FOXA2 were prominently bound at this region in islets (**Figure 3b**). FOXA2 loss-of-function causes CHI^23^, and NKX2-2 is part of a large repressor complex that regulates beta-cell specification^24^ and could therefore have a role in maintaining the repressive epigenetic state at the HK1 locus. Multiple members of the NFAT family are expressed in beta-cells (**Supplemental Figure 6**), with NFATC2 inducing beta-cell proliferation in human islets^25^. Whilst NFATC2 is expressed at low levels in adult beta-cells it has greater expression in pancreatic progenitors (**Supplemental Figure 6**), supporting HK1 repression as mediated by loss of NFAT binding in late stages of beta-cell development and proliferation. Analysis of snATAC-data from human islets revealed endocrine cells present in hormone-high and hormone-low states^16^, with the former associated with increased promoter accessibility over secreted endocrine genes and the latter over cell-cycle genes. Our critical region is accessible in the hormone-high state of alpha, beta, and delta-cells, and only remains accessible in beta-cells in the hormone low state (**Supplemental Figure 5a**). Therefore, accessibility may be maintained at the critical region during cell proliferation, by factors including NFATC2, thereby ensuring HK1 is repressed in beta-cells.

Discovering non-coding regulatory mutations in *HK1* enabled us to identify the first regulatory element critical for the selective silencing of an otherwise ubiquitously expressed gene within a single cell-type. It is interesting to note that interruption of transcription factor binding is implicated in each of the 9 distinct mutations and the locus is decorated by both active and repressive epigenetic marks suggesting that continual transcription factor binding is necessary to maintain repression in beta-cells. Future work should prioritise regulatory regions surrounding disallowed genes with similar epigenetic marks and to explore whether different regulatory mechanisms exist to silence disallowed genes within the beta-cell.

Over 60 beta-cell disallowed genes have been described, although the mechanism(s) controlling cell-specific silencing in humans have not been fully determined^26,27^. Linkage analysis of *HK1* to CHI^28^ and increased HK1 expression^29^ have been reported in several patients, but disease causality and genetic aetiology were not established. Novel promoter variants in the beta-cell disallowed gene, *SLC16A1* have also been reported in two families with exercise–induced hyperinsulinism. However, these variants increased transcription across cell types leading to the hypothesis that differences in the post-transcriptional regulation of mRNA across tissues could explain the beta-cell-specific phenotype^30^.

The identification of non-coding *HK1* mutations in patients with CHI represents one of the first discoveries of regulatory non-coding mutations affecting a gene in which coding mutations do not cause the same phenotype^31^. These findings highlight a role for undiscovered regulatory mutations causing disease through inappropriate expression of a normally functioning protein in a specific cell-type.

Mutations affecting the regulation of *HK1* cause CHI through its aberrant expression in beta-cells. These findings are important for future efforts to discover non-coding regulatory mutations as they establish a critical role for disallowed genes in Mendelian disease.

## Data Availability

All data produced in the present study are available upon reasonable request to the authors.

## Acknowledgements

SEF has a Sir Henry Dale Fellowship jointly funded by the Wellcome Trust and the Royal Society (105636/Z/14/Z). MaNW and MBJ are the recipients of an Independent Fellowship and TWL and NO a Lectureship from the Exeter Diabetes Centre of Excellence funded by Research England’s Expanding Excellence in England (E3) fund. EDF is the recipient of a Diabetes UK RD Lawrence fellowship (19/0005971). KAP has a Career Development fellowship funded by the Wellcome Trust (219606/Z/19/Z). IB and MJD were supported by the Northern Congenital Hyperinsulinism (NORCHI) charitable fund, by the Manchester Academic Health Sciences Centre and by The University of Manchester MRC Confidence in Concept (CiC) Award (MC_PC_18056). SJR and NGM are grateful to JDRF for funding (2-SRA-2018-474-S-B). Ivo Barić is member of the European Reference Network for Rare Hereditary Metabolic Disorders (MetabERN), Project ID No 739543.This research was performed with the support of the Network for Pancreatic Organ donors with Diabetes (nPOD; RRID: SCR_014641), a collaborative type 1 diabetes research project supported by JDRF (nPOD: 5-SRA-2018-557-Q-R) and The Leona M. & Harry B. Helmsley Charitable Trust (Grant#2018PG-T1D053, G-2108-04793). The content and views expressed are the responsibility of the authors and do not necessarily reflect the official view of nPOD. Organ Procurement Organizations (OPO) partnering with nPOD to provide research resources are listed at http://www.jdrfnpod.org/for-partners/npod-partners/. This research has been conducted using the UK Biobank Resource. This work was carried out under UK Biobank project number 9055 and 9072. Targeted next-generation sequencing to exclude known disease-causing mutations was funded by Congenital Hyperinsulinism International (a 501(c)3 organisation) for 20 individuals within the discovery cohort.

## Author Contributions

S.E.F. designed the study. A.D, S.E., I.B., S.E.F. and the International Congenital Hyperinsulinism Consortium recruited patients to the study. Ma.N.W*, M.B.J., J.A.L.H., R.V-H., S.E. and S.E.F. performed the molecular genetic analysis and the interpretation of resulting data. Ma.N.W. and Mi.N.W. performed the bioinformatic analysis, T.I.H., J.H., I.B., A.T.H. and S.E.F. analysed the clinical data. E.C. and M.J.D. performed the pathological analysis of the pancreatic tissue, J.R.H, C.S.F., R.W., N.G.M. and S.J.R. designed and performed the immunohistochemistry studies, N.D.L.O. designed and performed the epigenomic data analysis, Ma.N.W., N.D.L.O., J.R.H, M.B.J., T.W.L., E.D-F., K.A.P., A.T.H., M.J.D., S.J.R and S.E.F. prepared the draft manuscript. All authors contributed to the discussion of the results and to manuscript preparation.

## ONLINE METHODS

### Subjects

Congenital hyperinsulinism (CHI) was defined as an inappropriately high level of plasma insulin at the time of hypoglycaemia associated with inappropriately supressed ketones and free fatty acids presenting within the first 12 months of life. The definition of hypoglycaemia was based on the recommendations of the Pediatric Endocrine Society (blood glucose <2.8mmol/L in the presence of detectable insulin)^32^. Clinical details of the subjects are provided in **Supplemental Table 1**. Subjects with CHI were recruited by their clinicians for molecular genetic analysis to The Exeter Genomics Laboratory. Disease-causing mutations in the known CHI genes had been excluded by targeted-next generation sequencing in all cases^33^.

### Ethical considerations

This study was approved by the North Wales Research Ethics Committee (517/WA/0327) and was conducted in accordance with the Declaration of Helsinki, with all subjects or their parents providing informed consent for genetic testing. All tissue samples were studied with full ethics approval (West of Scotland Research Ethics Committee, reference: 20/WS/0074 IRAS project ID: 283620 or nPOD)^34^.

### Whole-genome sequencing

Whole-genome sequencing of DNA extracted from peripheral blood leukocytes was completed on 135 probands with CHI (n=3 with Illumina HiSeq 2500, n=69 with Illumina HiSeq X10, n=63 with BGISeq-500). The mean read depth across the whole genome was 36.9 (stddev 4.9). An additional 191 family members were also sequenced. The sequence data was aligned using BWA MEM 0.7.15, and processed using a pipeline based on the GATK best practices (Picard version 2.7.1, GATK version 3.7). Variants were annotated using Alamut batch standalone version 1.11 (Rouen, France).

CNVs were called across the genome by SavvyCNV^35^ using a bin size of 2Kbp. We also developed a new tool (FindLargeInsertSizes, available at https://github.com/rdemolgen/SavvySuite) to detect small (≥1Kbp) deletions, insertions, inversions, and translocations using read pair information. We used this tool to screen all whole genome sequenced samples for structural variants within *HK1* intron 2.

When a mutation was identified, Sanger sequencing (for point mutations/indels) or digital droplet PCR (for deletions) was performed on samples from each available family member to confirm genome sequencing results.

### Digital droplet PCR (ddPCR)

was performed on leukocyte DNA from patients 1 and 2.1 and their unaffected parents to confirm the results of the whole-genome sequencing deletion analysis. ddPCR was also performed on the affected twin brother of patient 2.1 and the 27 individuals with CHI of unknown cause who had undergone pancreatectomy.

Relevant primers (**Supplemental Table 3**) were used in reactions for droplet generation, PCR and detection using the Bio-Rad QX200 ddPCR EvaGreen system (Hercules, CA, USA) to search for copy number changes. Reactions were performed as per the manufacturer’s recommendations with an annealing/extension temperature during PCR of 59°C. All data were analysed using Bio-Rad QuantaSoft software.

### Sanger sequencing of the *HK1* regulatory element

Sanger sequencing was performed on DNA extracted from peripheral blood leukocytes from 27 individuals with CHI of unknown cause who had undergone pancreatectomy. Briefly, a 397bp genomic region (GRCh37/hg19, Chr10:71,108,536-71,108,932) within intron 2 of *HK1* (NM_033497) was amplified by PCR (primer sequences are listed in **Supplemental Table 3**). PCR products were sequenced on an ABI3730 capillary machine (Applied Biosystems, Warrington, UK) and analysed using Mutation Surveyor v3.24 software (SoftGenetics, State College, PA, USA). When a mutation was identified, samples from family members were tested to investigate co-segregation and microsatellite analysis using the PowerPlex kit (Promega, Southampton, UK) was performed to confirm family relationships.

### Histopathology

Formalin-fixed paraffin-embedded pancreatic tissue was available for immediate analysis from two individuals with *HK1* mutations age-matched controls (n=4) and age-matched tissues from patients with *ABCC8* mutations (n=2). Immunohistochemistry was performed as described previously on 5-mm-thick sections of tissue^36^. For high-content quantification of tissue, sections were first digitized using the 3DHistech Pannoramic 250 Flash II slide scanner (3DHISTECH, Budapest, Hungary) and quantified using QuPath^37^.

### Immunofluorescence

After dewaxing and rehydration, pancreatic samples from two *HK1* patients and 4 age-matched control tissues (patients with *ABCC8* mutations (n=2); non-hyperinsulinism donors (n=2)) were subjected to heat–induced epitope retrieval (HIER) in 10mM citrate pH6 buffer for 20 minutes. After blocking, the sections were probed in a sequential manner with rabbit monoclonal anti-hexokinase 1 (Abcam ab150423; 1/100 overnight); mouse monoclonal anti-glucagon (Abcam ab10988; 1/2000 for 1h) and guinea-pig anti-insulin (Agilent; C#IR002; 1/5 for 1h). The relevant antigen-antibody complexes were detected using secondary antibodies conjugated with fluorescent dyes (Alexa Fluor™ anti-mouse 555, anti-rabbit 488, anti-guinea pig 647) (Invitrogen, Paisley, U.K). Cell nuclei were stained with DAPI. After mounting, the sections were imaged via a Leica DMi8 confocal microscope (Leica Microsystems UK, Milton Keynes, UK) and the distribution of HK1, insulin and glucagon examined in multiple islets. In addition, sections were scanned at 40x magnification using an Akoya Biosciences Vectra® Polaris™ Automated Quantitative Pathology Imaging System. The quantification of the HK1 expression in islets was determined using the Random Forest Classifier Module (Version 3.2.1851.354), DenseNet AI V2 and HighPlex FL v4.04 modules included in Indica Labs HALO Image analysis platform (Version 3.2.1851.354). The Random Forest Classifier was used to identify the islets, then the DenseNet AI V2 module was used to identify beta and alpha cells within the islets. This enabled exclusion of HK1 positive non-islet cells such as red blood cells, stellate cells and endothelial cells which are frequently found in proximity to the islet. The median fluorescence intensity (MFI) of HK1 expression in beta and alpha cells were then calculated using the HighPlex FL v4.04 module. This pipeline was applied to all corresponding tissue sections. GraphPad Prism V9.2.0 (332) was used to demonstrate the median and IQR of beta and alpha cells MFI in each of the donors assessed.

### Epigenomic analysis

#### Public Bulk ChIP and ATAC-seq

For ChIP-seq and ATAC-seq datasets reads were downloaded from accessions provided in (**Supplemental Table 4**), and then aligned with Bowtie2^38^ to the GRC37/hg19 genome with Bowtie2 v2.3.5.1 with default parameters for single end reads and with additional options “- I 0 -X 1000 --no-discordant --no-mixed” for paired-end reads. Alignments were filtered for those with mapping quality > 30 and then reads with identical aligning coordinates were treated as duplicates and collapsed to a single alignment. All data is visualised as reads per million using https://github.com/owensnick/GenomeFragments.jl. For human islet single-nuclei ATAC-seq data^16^ (GSE160472), reads from the authors’ combinatorial barcode approach were aligned as above and separated into cell types using author provided cluster labels: vhttps://github.com/kjgaulton/pipelines/tree/master/islet_snATAC_pipeline.

Human islet Hi-C data was obtained from experiment accession TSTSR043623 and file accession DFF064KIG (.hic file) and TSTFF938730 (bedpe file)^17^. The .hic file from experiment was downloaded and converted to .cool format using hic2cool https://github.com/4dn-dcic/hic2cool with default parameters. Contact matrices were obtained using cooler^39^ at 5kb resolution using KR balance and log-transformed for visualisation.

#### scRNA-seq Expression Datasets

scRNA-seq data collected over a time course of pancreatic differentiation^18^ projected onto a differentiation pseudotime was obtained from the reference’s supplementary table. We identity consistent temporal trends using Gaussian Process (GP) regression, following the approach we have previously applied^40^. Briefly, to stabilise variance we transform data log(αy+ β) where y is the gene expression for a given gene and α = 100, β = 1, then perform GP regression using Matern52 kernel and invert the transformation to report GP median and 95% confidence intervals. All GP regression was performed with GaussianProcesses.jl (https://github.com/STOR-i/GaussianProcesses.jl; https://arxiv.org/abs/1812.09064).

For scRNA-seq data in human islets^41^ accession GSE101207 we gene counts per cell for the size healthy donors and normalised by depth per cell. In **Supplemental Figures 3, 6 and 7** we show boxplots of normalised counts for each gene over healthy donors.

Finally, we obtained data on gene expression during beta-cell maturation from the Broad Single Cell portal^19^. In **Supplemental Figures 3, 6 and 7** we show mean normalised counts for each gene over annotated cell types.

### Motif analysis

To assess motifs maximally disrupted by the mutations, we took all motifs in the non-redundant JASPAR database^42^ and found the maximal scoring match that spanned the position of each mutation for both reference and mutated sequences using https://github.com/exeter-tfs/MotifScanner.jl. Motif scores are likelihood-ratio scores of motif PWM over a background of the A,C,G,T frequencies in the hg19 genome. We normalised all motif scores, by the maximal score for each PWM and to both primary and secondary candidate motifs that may be disrupted by the mutations. We considered two tiers; Tier 1 having normalised motif score ≥ 0.6, and Tier 2 having 0.45 ≤ normalised motif score < 0.45. For each tier we calculated a disruption score by subtracting the mutated sequence score from the reference sequence score and ranked these across all motifs. To report a single disrupted motif family, we grouped multiple motifs with overlapping alignments by family by removing any trailing numbers from the gene symbol (e.g. NKX2-2 → NKX2, HIC2 → HIC) and all FOX factors we grouped in the FOX family. Tier 1 disrupted motif families include NFAT, NKX2 and FOX (**Figure 3e**). One has two *de novo* mutations, the leftmost is shared with other patients and is a candidate for NFAT disruption, and the rightmost interrupts a HIC family motif (**Supplemental Figure 7a**). HIC family members include HIC1 and HIC2, with HIC2 the most greatly expressed in beta-cells (**Supplemental Figure 7b**). Interestingly, HIC2 is a transactivator of SIRT1^43^ and the loss of SIRT1 impairs glucose sensing in beta-cells in mice^44^. Tier 2 disrupted motif families include TEAD and SMAD (**Supplemental Figure 7c**). The TEAD family motif shares TTCA consensus with the NFAT family and is an alternative candidate to NFAT, TEAD1 is expressed in beta-cells (**Supplemental Figure 7d**) and plays a critical role in pancreatic progenitors^21^, however it should be noted that TEAD1 does not bind the critical region in pancreatic progenitors when the region is bound by FOXA2 (**Figure 3b**). Multiple members of the SMAD family are expressed in beta-cells (**Supplemental Figure 7e**), SMAD factors are signal transducers of TGF-beta signalling and play an important role in beta-cell development, function, and proliferation^45^.

