## Supplemental Data for "A novel disease mechanism leading to the expression of a disallowed gene in the pancreatic beta-cell identified by non-coding, regulatory mutations controlling *HK1*"

**Supplemental Table 1:** Summarised clinical characteristics for 162 probands diagnosed with congenital hyperinsulinism screened for mutations in the *HK1* regulatory region by whole-genome sequencing (n=135) or Sanger sequencing and ddPCR (n=27). Median values (IQR) provided**.**

| | **Summary data** |
|---|---|
| **Female sex** | 80 (49.1%) |
| **Birth weight SDS** | 0.10<br>(-1.16 – 1.14) |
| **Age at onset (days)** | 56<br>(7 – 168) |
| **Blood glucose at presentation (mmol/l)** | 1.6<br>(1.2 – 2.1) |
| **Insulin at presentation (pmol/l)** | 48<br>(25 – 100) |
| **Pancreatectomy performed** | 34 (20.8%) |

**Supplemental Table 2:** Summarised clinical characteristics for 17 individuals from 14 families with *HK1* mutations. Median values (IQR) provided**.**

| | **Summary data** |
|---|---|
| **Sex (female)** | 9 (53%) |
| **Birth weight (SDS)** | 0.61<br>(0.10-1.84) |
| **Age at onset** | Birth<br>(0-14 days) |
| **Blood glucose at presentation (mmol/l)** | 1.6<br>(1.2-2.1) |
| **Insulin during hypoglycaemia (pmol/l)** | 163<br>(95-326) |
| **Pancreatic resection performed** | 5/17 (29%) |

**Supplemental Table 3**: Primer sequences used for **a)** ddPCR to confirm the deletions identified by whole-genome sequencing in two individuals and an affected twin **b)** ddPCR to screen for deletions in the replication cohort within the 151bp region harbouring *de novo* single nucleotide variants **c)** Sanger sequencing of the *HK1* element

**a)**

| Primer | Sequence (5' – 3') | Region amplified (Hg19) |
|---|---|---|
| HK1_A_Forward | tccctgcagtaatcagaatctgg | Chr10:71105908-71106051 |
| HK1_A_Reverse | agcgccaatttatccctccg | |
| HK1_B_Forward | agggtctgcgtggtctttatg | Chr10:71107602-71107732 |
| HK1_B_Reverse | ggaaacccaaaatacttcacccc | |
| HK1_C_Forward | ctcttacagtggcagggacg | Chr10:71109774-71109906 |
| HK1_C_Reverse | aagccaatcacagccctctc | |
| HK1_D_Forward | gtgattggtacagaaagactcagtg | Chr10:71111625-71111726 |
| HK1_D_Reverse | cagtatgaagtagctcctcgcag | |

**b)**

| Primer | Sequence (5'-3') | Region amplified (Hg19) |
|---|---|---|
| GCK_exon_1_Foward | TCCACTTCAGAAGCCTACTG | Chr7:44228423-44228617 |
| GCK_exon_1_Reverse | TCAGATTCTGAGGCTCAAAC | |
| HK1_E_Forward | caggctggtactcgagacac | Chr10:71108653-71108835 |
| HK1_E_Reverse | tgcctgtgacctttcctcac | |
| HK1_F_Forward | aggcagagtttttgcttgcc | Chr10:71108620-71108804 |
| HK1_F_Reverse | ccactgaagctgggagaacc | |

**c)**

| | **Sequence (5'-3')** |
|---|---|
| **Forward** | AGCCTGGGCAACAGAAAC |
| **Reverse** | GCTACAAGCTCAGCCTCTTTC |

**Supplemental Table 4:** Table describing the primary and secondary candidate disrupted transcription factor motifs by each mutation.

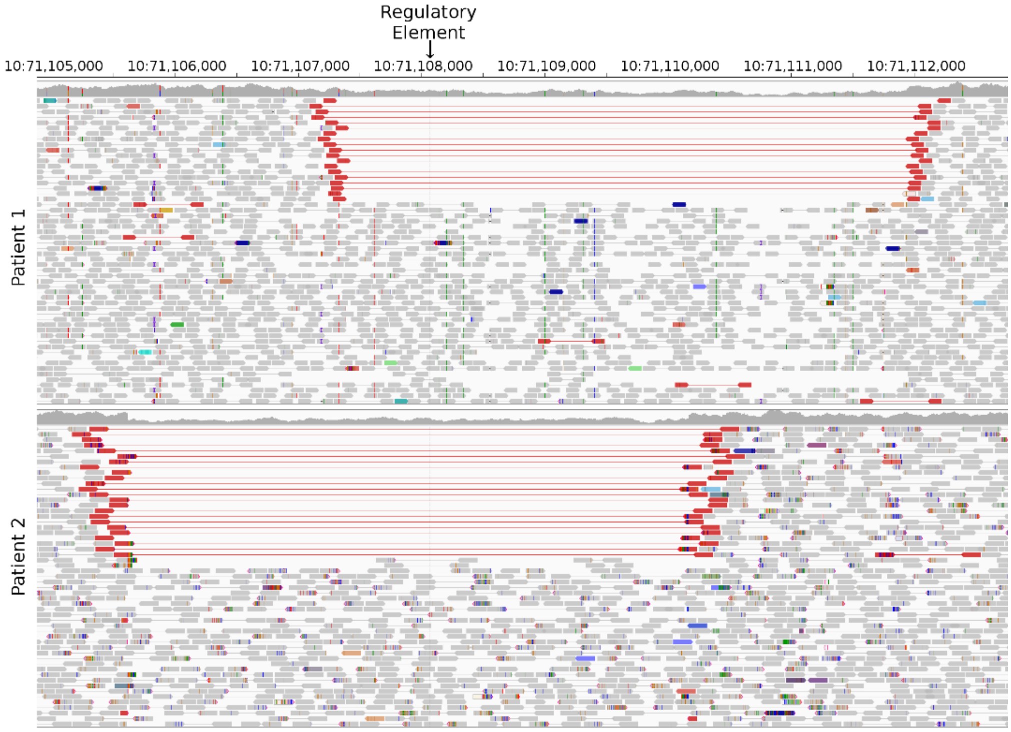


**Supplemental Figure 1: Integrative Genomics Viewer (IGV) screenshot of the ~4.5Kb deleted regions encompassing the regulatory element in intron 2 of *HK1* in two patients.** Both deletions are heterozygous, so reads are present in the deleted regions from the other haplotype, although the read depth is reduced. Reads highlighted in red have a long insert size. Both features are consistent with a deletion. The deleted region in one patient has detectable break points, as shown by the discontinuity in read depth and the reads clipped at the deletion boundary. In the second patient, the breakpoints do not result in read depth discontinuity or clipped reads as the deletion occurs between two matching repeating sequences.

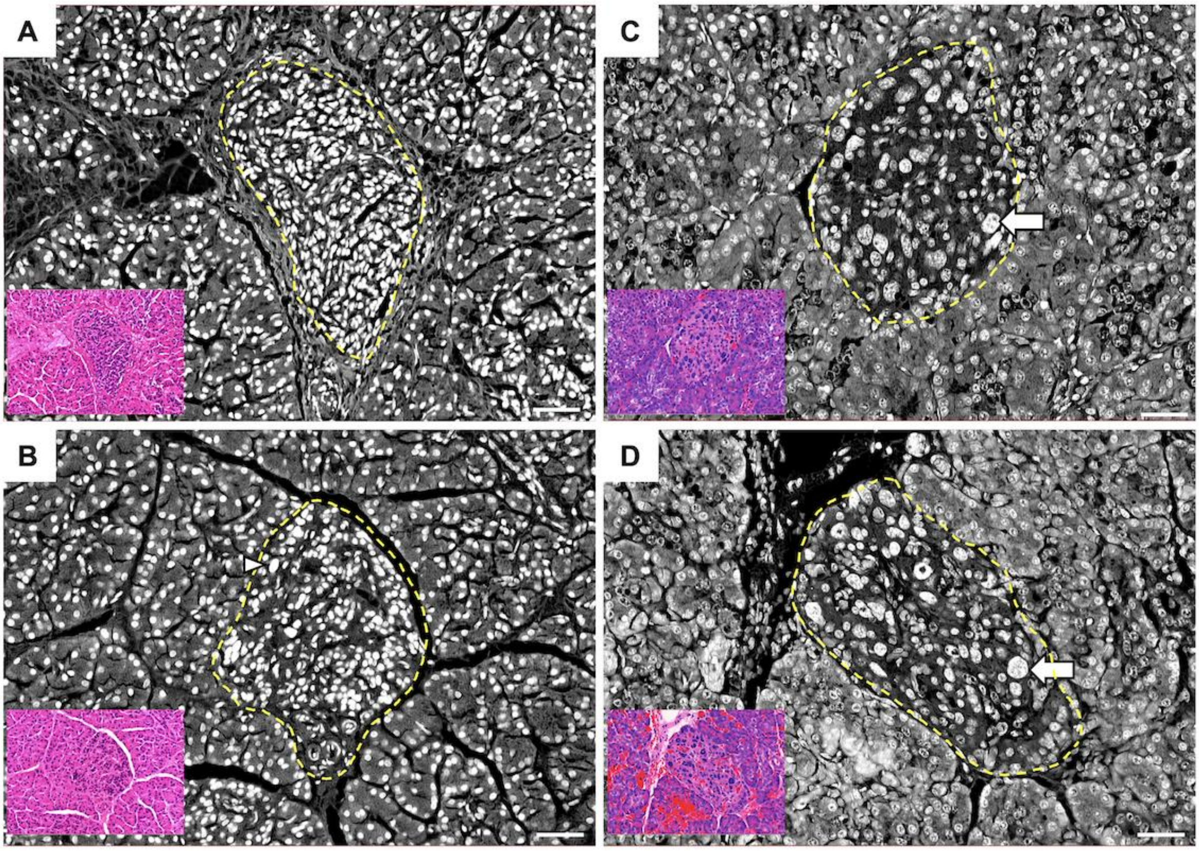


**Supplemental Figure 2. Histological appearance of islets from *HK1*-CHI and *ABCC8*-HI tissue**. Panels **A** and **B** illustrate islets from two *HK1* CHI patients in comparison to age-matched tissue from *ABCC8* CHI patients with diffuse pancreatic disease **(C and D).** The yellow dashed line delineates a single islet in each sample. Nuclear enlargement (B, arrowhead) and nucleomegaly (C, D arrows) was virtually undetected in control islets (n=12/72,145 islet cells) but found in 0.8% of HK1 islets (n=374/46,885 cells) and 4.9% of *ABCC8* islet cells (n=70,929). Inserts illustrate the corresponding haematoxylin and eosin images. Scale bar 50um

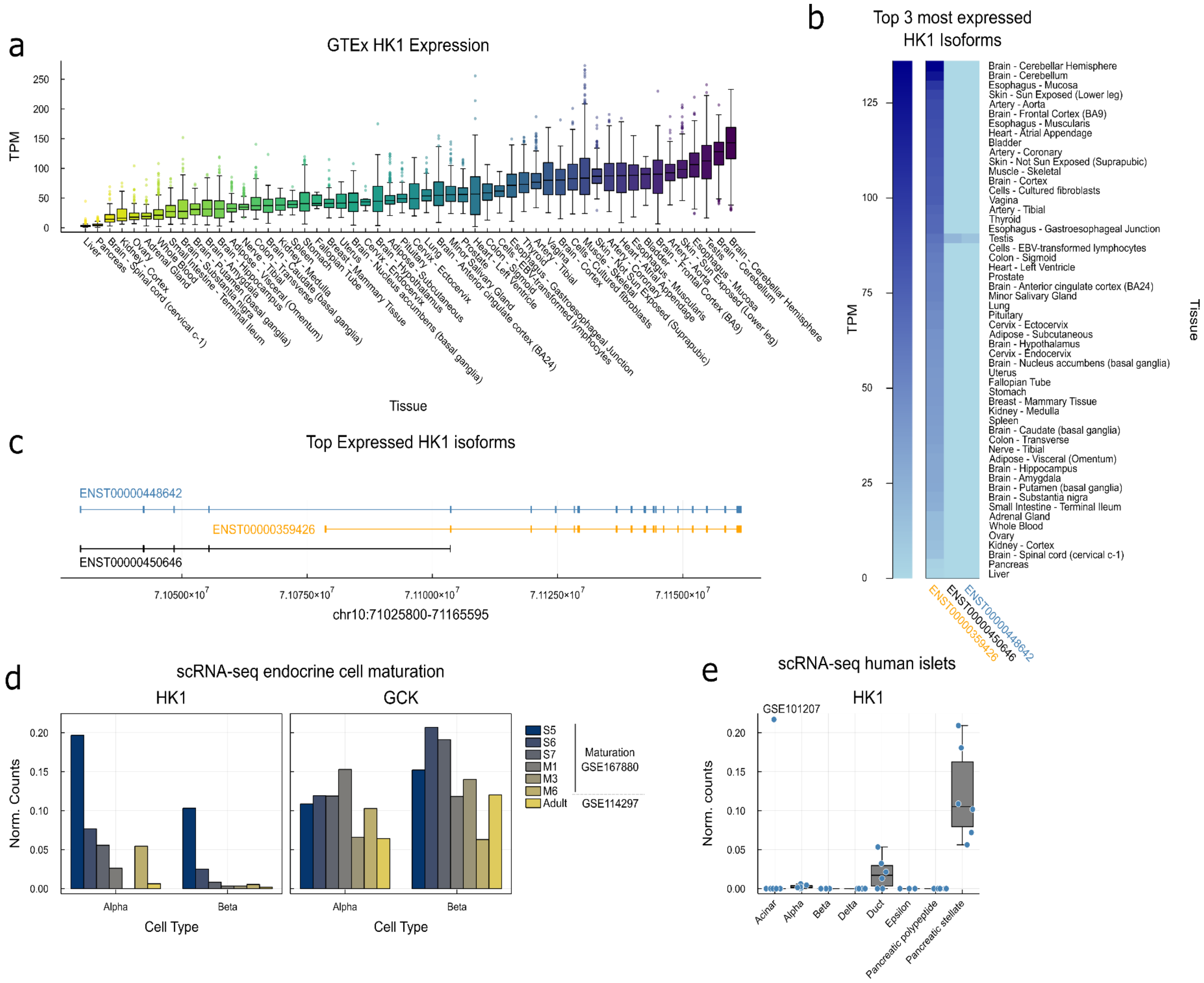


**Supplemental Figure 3: HK1 expression in GTEx a)** Boxplots describing gene level HK1 expression in Genotype-Tissue Expression GTEx project ordered by expression with least expression in liver and pancreas. Data shows sum of GTEx v8 isoform level data calculated with RSEM. The GTEx Project was supported by the Common Fund of the Office of the Director of the National Institutes of Health, and by NCI, NHGRI, NHLBI, NIDA, NIMH, and NINDS. The data used for the analyses described in this manuscript were obtained from the GTEx Portal on 08/26/21. **b)** Isoform level HK1 expression in GTEX tissues as a), top 3 most expressed isoforms are given. Long HK1 isoform only expressed in testis, short isoform ubiquitously expressed across all tissues. **c)** Transcript models of top three HK1 isoforms given in b). **d)** Expression of HK1 and GCK for comparison in scRNA-seq across alpha and beta-cell maturation, all data quantified by (GSE167880)[1], which quantified their own data over maturation and adult endocrine cells (GSE114297)[2]. **e)** HK1 expression in human islet cell types by scRNA-seq (GSE101207)[3]. Data points from independent human donors, boxplots describing median and IQR. HK1 expression only in duct and pancreatic stellate cells, expression absent in endocrine cell types.

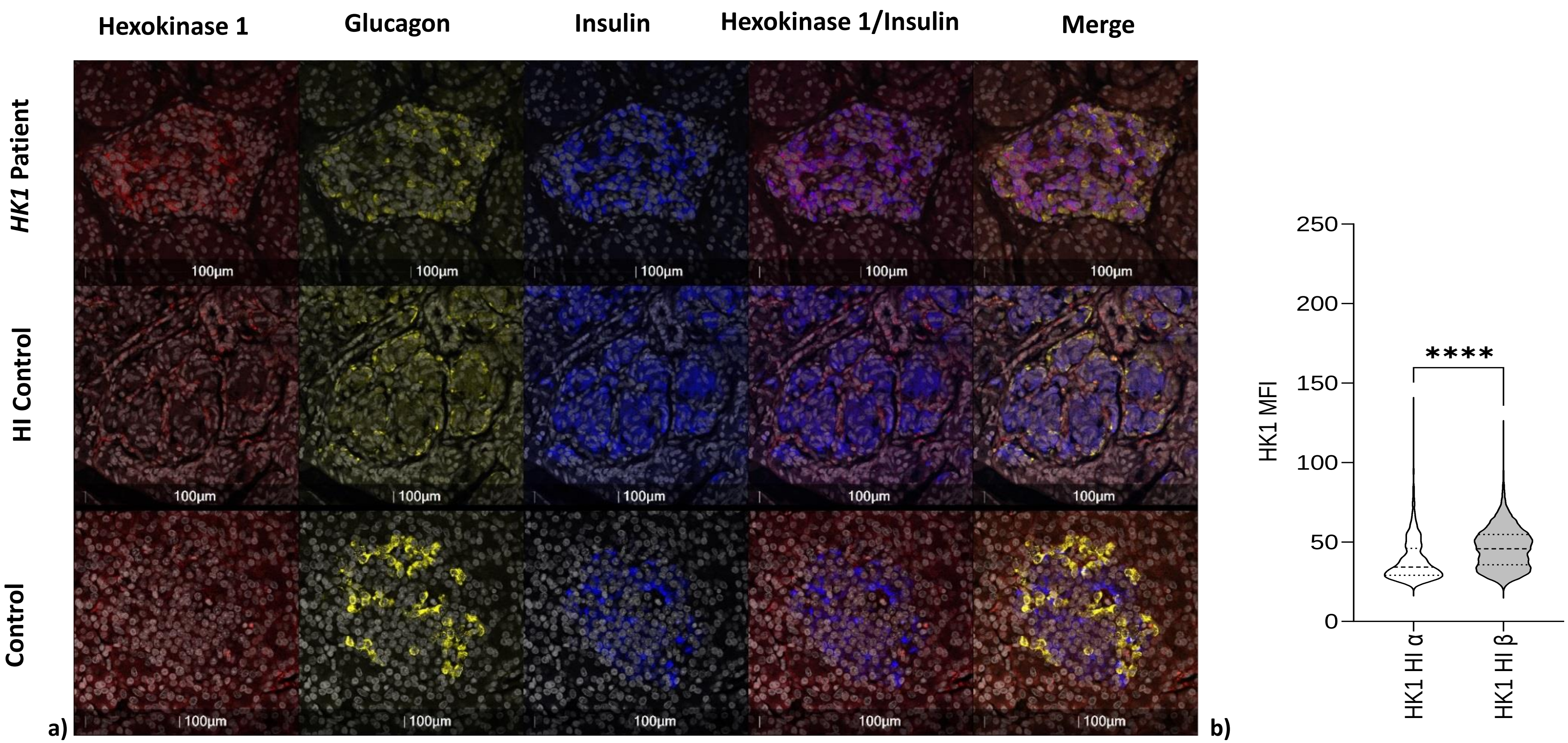


**Supplementary Figure 4: HK1 expression is present in the beta-cells of donors with *HK1*-CHI, but not *ABCC8*-Hyperinsulinism or Non-HI controls. a)** Staining of HK1 (red); glucagon (yellow); insulin (blue) and DAPI (white) in pancreatic tissue resected from one patient with a *HK1* mutation. HK1 co-localises with insulin (purple staining), but not with glucagon. Staining in tissue resected from an individual with *ABCC8*-Hyperinsulinism and a Non-HI control shows no expression of HK1 in either alpha (glucagon-positive) or beta (insulin-positive cells). Scale bar – 100µm. **b)** HK1 expression (Median fluorescence intensity (MFI)) is higher in the *HK1* HI donor beta-cells (n=2 donors; 17015 beta-cells), when compared to alpha-cells (n=3034). Two-tailed Mann-Whitney (U=16323914; ****$p<0.0001$). This suggests the impact of the *HK1* mutation is beta-cell specific.

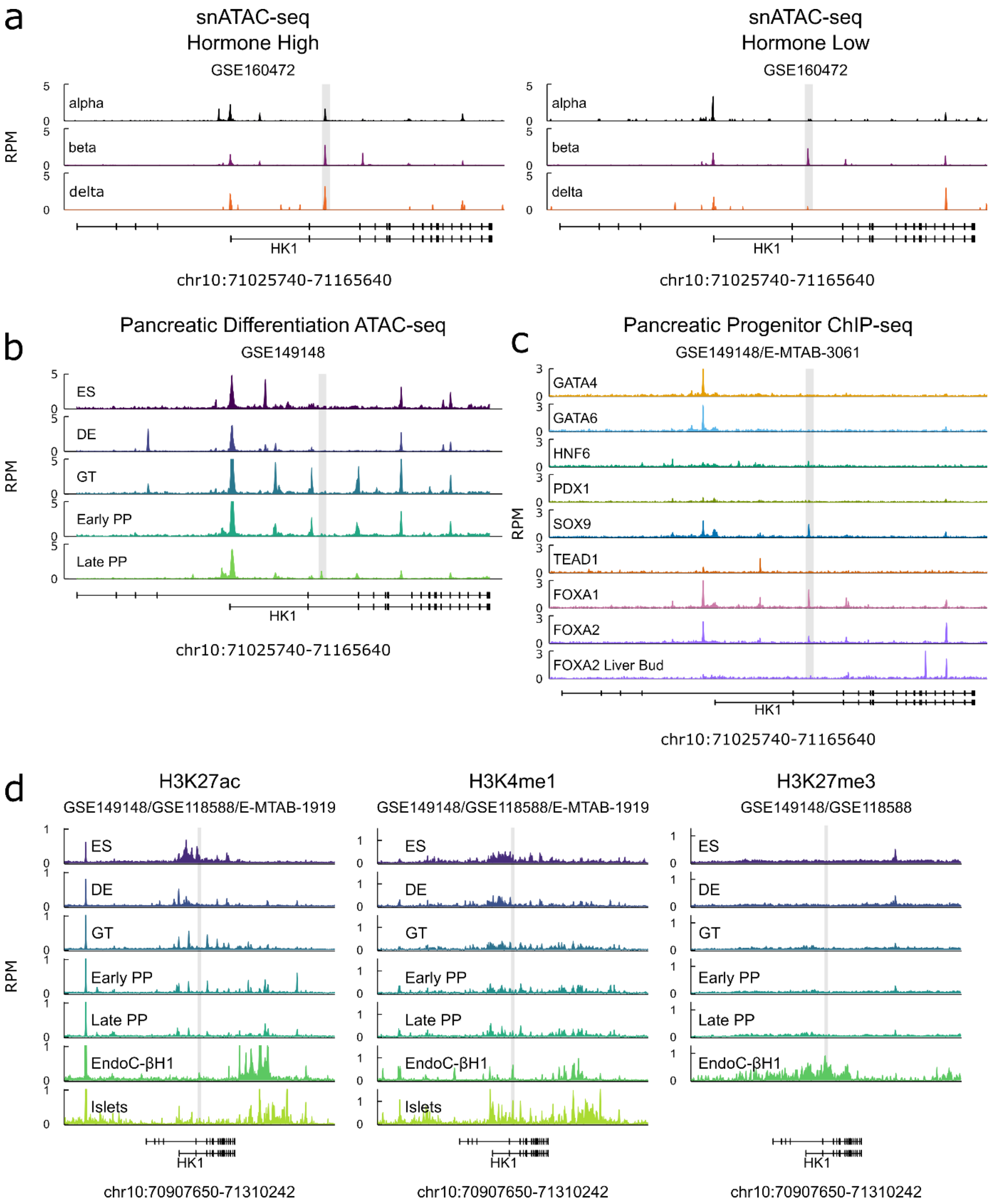


**Supplemental Figure 5: Chromatin accessibility, Transcription Factor binding and Epigenetic modifications at the *HK1* locus over pancreatic differentiation a)** snATAC-seq data in human islets showing mean chromatin accessibility over nuclei assigned to alpha, beta and delta cell clusters in hormone high and hormone low conditions (GSE160472)[4]. Critical region containing mutations encompassed within grey box. Alpha, beta and delta cells each show accessibility in hormone high state, whereas only beta-cells retain accessibility in hormone low cells. Hormone high and low states characterised by level of accessibility over endocrine cell secreted hormone promoter, alpha – GCG, beta –

INS, delta – SST. Data show in reads per million (RPM). **b)** Chromatin accessibility of pancreatic differentiation assessed by ATAC-seq (GSE149148)[5], wider locus view of same data shown in **Figure 3c**. Critical region containing mutations encompassed within grey box. Stages: ES – embryonic stem cell, DE – definitive endoderm, Early/Late PP – pancreatic progenitor. **c)** Transcription factor binding in pancreatic progenitors (GSE149148)[5] and in Carnegie Stage 16-18 liver bud for FOXA2 (E-MTAB-3061)[6], data shown as a) and b). Liver bud data reveals that whilst FOXA2 binds critical region in pancreatic progenitors and human islets (**Figure 3b**), it does not bind in liver bud, suggesting that critical region encodes pancreas specific regulation of *HK1*. **d)** Epigenetic modifications over *HK1* locus, shown are active marks H3K27ac and H3K4me1 and repressive mark H3K27me3 over in vitro pancreatic cell differentiation (GSE149148)[5] stages as in b; in vitro beta-cell line EndoC-BH1 (GSE118588)[5] and human islets (E-MTAB-1919)[7]. Grey box encompasses critical region containing mutations. Data reveals that H3K27ac broadly marks short isoform promoter (**Supplemental Figure 3**) at ES cells stage is reduced by DE stage, whilst focal mark over regulatory region is present in EndoC-BH1 and human islets. Similarly, H3K4me1 shows string focal enrichment over regulatory region in EndoC-BH1 and islets. Finally, repressive mark H3K27me3 is broadly absent over the locus over the course of pancreatic cell differentiation but broadly marks region in mature Endoc-BH1 cells.

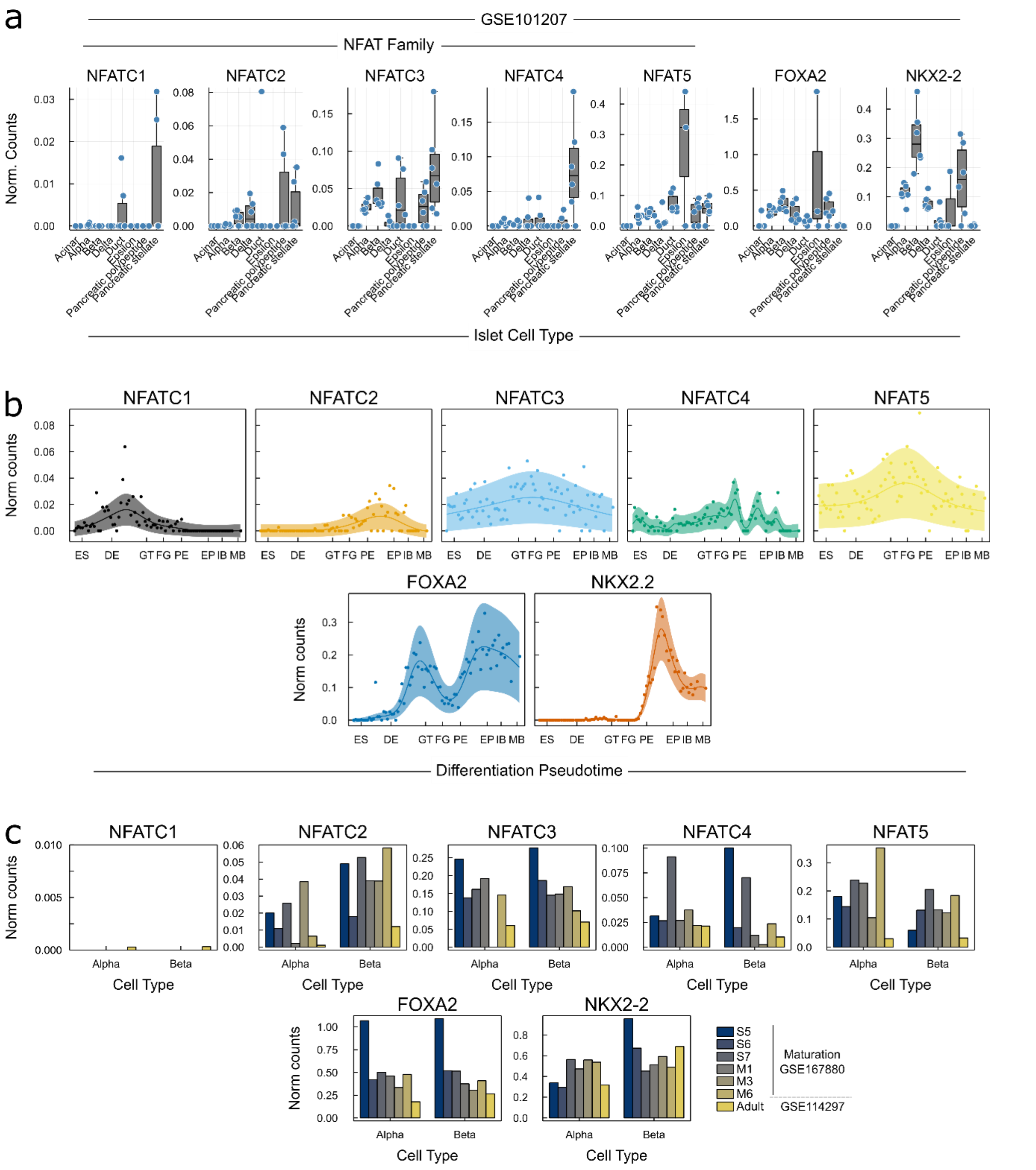


**Supplemental Figure 6: Expression of family members of disrupted transcription factor binding motifs in critical region**: NFAT family, NKX2-2 and FOXA2: **a)** In single-cell RNA-seq in human islets (GSE101207)[3], boxplots of mean normalised counts from independent islet donors in cells assigned to islet cell type clusters (data shown as in **Supplemental Figure 3e**). **b)** Over the course of pancreatic differentiation from embryonic stem cells to maturing beta-cells, expression data over a beta-cell differentiation psuedotime[8], Gaussian process regression median and 95% CI shown. ES – embryonic stem cell, DE – definitive endoderm, GT – gut tube, FG – foregut, PE – pancreatic endoderm, EP – endocrine precursors, IB – immature beta-cells, MB – maturing beta-cells. **c)** scRNA-seq across alpha and beta-cell maturation, all data quantified by (GSE167880)[1], which quantified their own data over maturation and adult endocrine cells (GSE114297)[2].

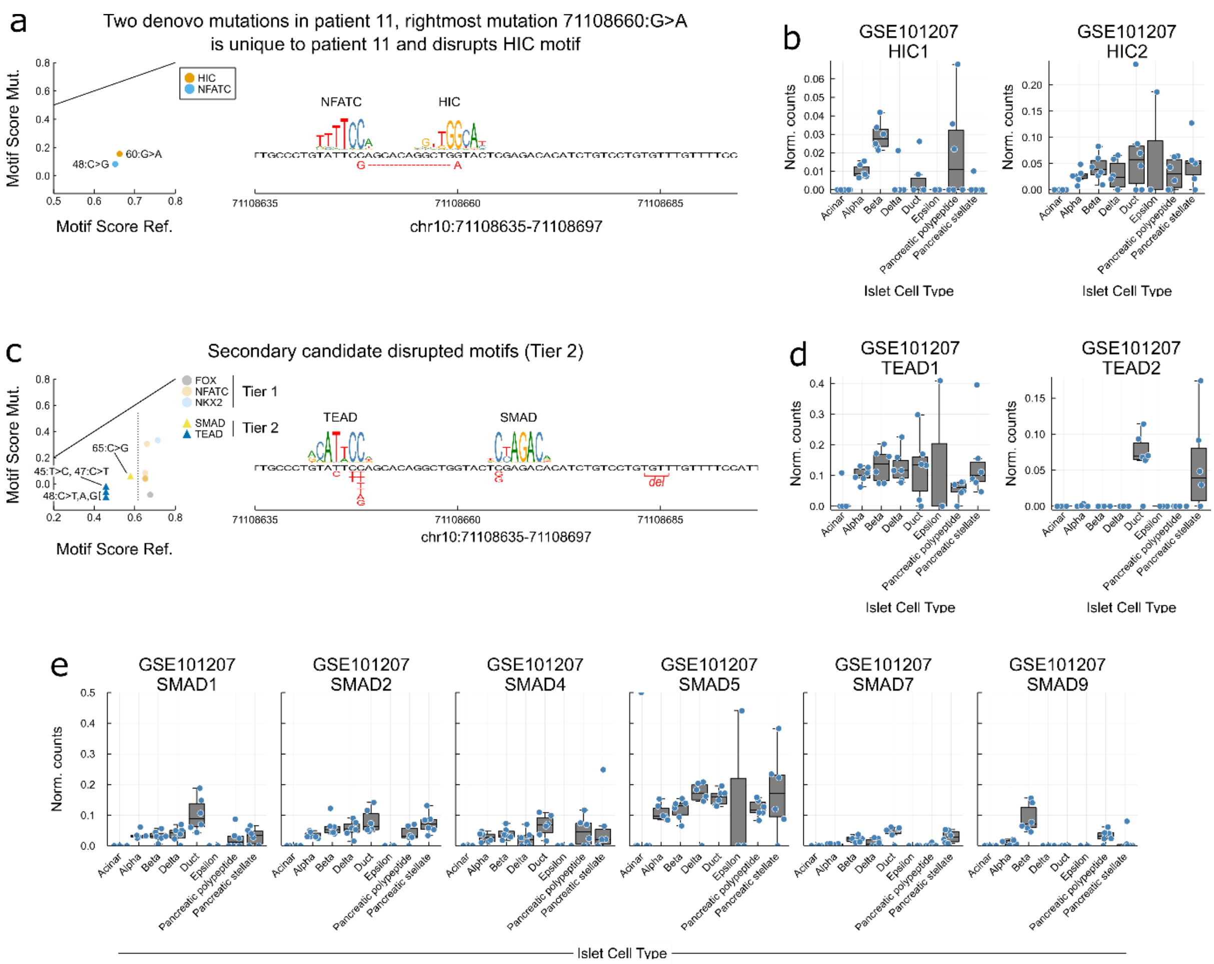


**Supplemental Figure 7: Additional transcription factor motifs disrupted by mutations**
**a-b)** Sanger sequencing identified 2 *de novo* variants 12 base pairs apart (g.71,108,648C>G and g.71,108,660G>A) in one patient. It is not known whether these variants are *in cis* or *in trans*. g.71,108,648 is affected by 3 different *de novo* substitutions in 6 individuals and resides within a predicted NFATC binding motif. Two further *de novo* mutations identified in 3 patients are also predicted to impact on NFATC binding (**Figure 3**). In contrast, further variants affecting g.71,108,660 have not been identified, this suggests that the g.71,108,648C>G is causative of the disease in this patient. Nevertheless, it is of note that g.71,108,660G>A is predicted to affect a HIC family motif **a)** and human islet scRNA-seq (GSE101207)[3] determines HIC2 is expressed in beta-cells **b)** Interestingly, HIC2 is a transactivator of SIRT1[9] and the loss of SIRT1 impairs glucose sensing in beta-cells in mice[10]. **c-e)** Tier 2, secondary candidate motifs for disruption have lower normalised motif scores than Tier 1 matches. **c)** Two additional motif families are implicated TEAD and SMAD. TEAD shares partial consensus with NFAT (TTCCA) and is alternative candidate to NFAT. TEAD1 but not TEAD2 is expressed in beta-cells **e**) and TEAD1 plays a critical role in pancreatic progenitors[6], however it should be noted that TEAD1 does not bind the critical region in pancreatic progenitors when the region is bound by FOXA2 (**Figure 3b**). The SMAD family are signal transducers of TGF-beta signalling and play an important role in beta-cell development, function, and proliferation[11].

**Supplemental Note**

**The International Congenital Hyperinsulinism Consortium**

Ivo Barić[1], Liat de Vries[2,3], Samar S. Hassan[4], Khadija Nuzhat Humayun[5], Floris Levy-Khademi[6], Catarina Limbert[7], Birgit Rami-Merhar[8], Verónica Mericq[9], Kristen Neville[10], Yasmine Ouarezki[11], Ana Tangari[12], Charles Verge[10], Esko Wiltshire[13]

1. Division for Genetics and Metabolic Diseases, Department of Pediatrics University Hospital Center Zagreb and University of Zagreb, School of Medicine, Kišpatićeva 12, 10000 Zagreb, Croatia
2. The Jesse Z and Sara Lea Shafer Institute for Endocrinology and Diabetes, National Center for Childhood Diabetes, Schneider Children's Medical Center of Israel, Petach Tikva, Israel
3. Sackler Faculty of Medicine, Tel Aviv University, Tel Aviv, Israel
4. Department of Pediatric Endocrinology, Gaafar Ibn Auf Pediatric Tertiary Hospital, Khartoum, Sudan.
5. Department of Paediatrics and Child Health, Aga Khan University, Karachi, Pakistan
6. Division of Pediatric Endocrinology, Shaare Zedek Medical Center, Faculty of medicine, Hebrew University, Jerusalem, Israel
7. Unit for Paediatric Endocrinology and Diabetes, Centro Hospitalar e Universitário de Lisboa Central, Hospital Dona Estefania, Lisbon, Portugal
8. Medical University of Vienna, Department of Pediatric and Adolescent Medicine, Vienna, Austria
9. Department of Pediatrics, Institute of Maternal and Child Research, Clinica Las Condes, Santiago, Chile
10. Endocrinology, Sydney Children's Hospital, Randwick, Australia and School of Women's and Children's Health, UNSW, Australia
11. EPH Hassan Badi, Algiers, Algeria
12. Saredo, Sanatorio Güemes, Pediatric Endocrinology Unit, Buenos Aires, Argentina
13. Department of Paediatrics and Child Health, University of Otago Wellington, Wellington, New Zealand
